# Population-based Characterization of PTEN Hamartoma Tumor Syndrome

**DOI:** 10.64898/2025.12.19.25342661

**Authors:** Gideon Idumah, Chloe Bautista, Lin Li, Lamis Yehia, Ying Ni

**Affiliations:** Cancer Sciences, Cleveland Clinic Research, Cleveland Clinic, Cleveland, Ohio, USA; Genomic Medicine Institute, Cleveland Clinic Research, Cleveland Clinic, Cleveland, Ohio, USA; Epilepsy Center, Neurological Institute, Cleveland Clinic, Cleveland, Ohio, USA; School of Medicine, Case Western Reserve University, Cleveland, Ohio, USA; Cleveland Clinic Lerner College of Medicine, Case Western Reserve University, Cleveland, Ohio, USA; Case Comprehensive Cancer Center, Case Western Reserve University, Cleveland, OH, USA

**Author notes:** **Corresponding Authors:** Ying Ni, PhD Department of Cancer Sciences, Cleveland Clinic Research 9500 Euclid Avenue NA20, Cleveland, OH 44195, USA, Lamis Yehia, PhD Epilepsy Center, Neurological Institute 9500 Euclid Avenue NA20, Cleveland, OH 44195, USA.

## Abstract

PTEN hamartoma tumor syndrome (PHTS) is a cancer predisposition disorder caused by germline *PTEN* variants, yet its full clinical spectrum remains poorly defined due to reliance on highly selected cohorts. Accordingly, PHTS is underrecognized and its prevalence underestimated. Leveraging genomic and electronic health record data from 414,830 participants in the All of Us (AoU) Research Program, we identified 55 individuals with pathogenic or likely pathogenic *PTEN* variants, the majority of whom lacked a prior PHTS diagnosis, underscoring underrecognition in the general population. PHTS affects ∼1/7500 individuals in this US cohort, which is about 26-folds higher than historical estimates for *PTEN*-related disorder. Compared with carriers of other cancer-related gene variants and noncarriers, *PTEN* variant carriers exhibited the highest cancer prevalence and significantly younger ages at first cancer diagnosis. Phenotype enrichment revealed expected overgrowth-related features as well as previously unreported associations, including adenotonsillar hypertrophy, sleep apnea, acanthosis nigricans, and extreme obesity, suggesting broader systemic involvement than classically appreciated. Variant spectra were consistent across the population-based and clinically-ascertained PHTS cohorts. These findings demonstrate that PHTS is more prevalent, more heterogeneous, and more often undiagnosed than current clinical practice reflects, emphasizing the value of population-scale genomics for comprehensive characterization and earlier detection of PHTS.

## Introduction

PTEN hamartoma tumor syndrome (OMIM 158350) is a cancer predisposition disorder caused by germline variants in the tumor suppressor gene *PTEN* (OMIM 601728).^1, 2^ PHTS encompasses several genetically related conditions, including Cowden syndrome (CS), Bannayan-Riley-Ruvalcaba syndrome (BRRS), and other *PTEN*-related disorders.^1, 2^ Germline *PTEN* variants are associated with increased lifetime risks of breast, thyroid, endometrial, kidney, and colon cancers, as well as melanoma.^3, 4, 5, 6, 7^ Individuals with PHTS also have notably elevated lifetime risks of second primary malignant neoplasms (SMN) compared with the general population.^8, 9^ Intriguingly, although considered a classical tumor suppressor gene, germline pathogenic *PTEN* variants are considered one of the most common genetic causes of monogenic autism spectrum disorder (ASD).^10, 11, 12^ As such, there is a well-established bi-modal distribution of PHTS phenotypes, whereby childhood diagnoses are enriched for ASD and/or other neurodevelopmental disorders (NDD), whereas adult diagnoses are enriched with cancer and overgrowth related phenotypes. Early identification and characterization of the natural history of PHTS are important, as prompt recognition facilitates gene-informed medical management, including proactive high-risk cancer surveillance and addressing the associated neurodevelopmental features.

Estimating the prevalence of PHTS has historically been challenging, mainly due to the wide phenotypic and genotypic variability, as well as the association with clinical features that are relatively common in the general population. Hence, many individuals remain undiagnosed, resulting in underestimated prevalence.^2^ A recent study evaluated the prevalence of PHTS in two large research cohorts, suggesting that PHTS may be 10-20 times more common than earlier estimates.^13, 14^ Relatedly, studies characterizing *PTEN* genotype and PHTS-related phenotypes have focused on subsets of individuals from specialized centers, such as our PTEN Multidisciplinary Clinic and Center of Excellence at the Cleveland Clinic, or centers focused on cancer care. Therefore, there are many individuals in the population who remain undiagnosed, and we posit that these individuals may have a different phenotypic spectrum than individuals presenting to specialized clinics due to a diagnosis with or a family history of cancer and/or NDD.

In this study, we leveraged the All of Us (AoU) research program to characterize phenotypes and genotypes of an unselected series of adults with *PTEN*-related syndromes. Additionally, we compared the phenotypic spectrum to that from individuals with pathogenic and/or likely pathogenic (P/LP) variants in other cancer predisposition genes from AoU, and to individuals from the Cleveland Clinic PTEN Multidisciplinary Clinic. Subsequently, we compared the *PTEN* genotype spectrum between AoU participants and those from the Cleveland Clinic PTEN Multidisciplinary Clinic. This effort led to the characterization of *PTEN*-related disorders in a population setting, thus optimizing the identification of PHTS, particularly in individuals unaffected by cancer and/or NDD.

## Results

### Identification of *PTEN*-related Disorders in All of Us Participants

For the purposes of phenotype-genotype characterization, we identified a total of 286,362 participants with both srWGS and phenotype data (**Fig. 1A**). Of the 353,834 participants with phenotype data, 25 individuals were reported to have PHTS, with only a subset reported to have germline P/LP *PTEN* variants. Relatedly, of 414,830 participants with srWGS, 55 individuals were reported to harbor germline P/LP *PTEN* variants (including two individuals with germline structural variants). Of the 46 individuals with EHR data, 37 lacked a formal PHTS diagnosis, suggesting that a subset of individuals with germline P/LP *PTEN* variants remain undiagnosed. A representation of a broad category of conditions grouped by 22 ICD-10 code chapters reveals an overrepresentation of phenotypes associated with the musculoskeletal system and connective tissues, in addition to endocrine, nutritional, and metabolic phenotypes (**Fig. 1B**). Notably, we did not identify any participant with reported neurodevelopmental disorders, due to limited information about these phenotypes in the AoU dataset.

**Figure 1:**
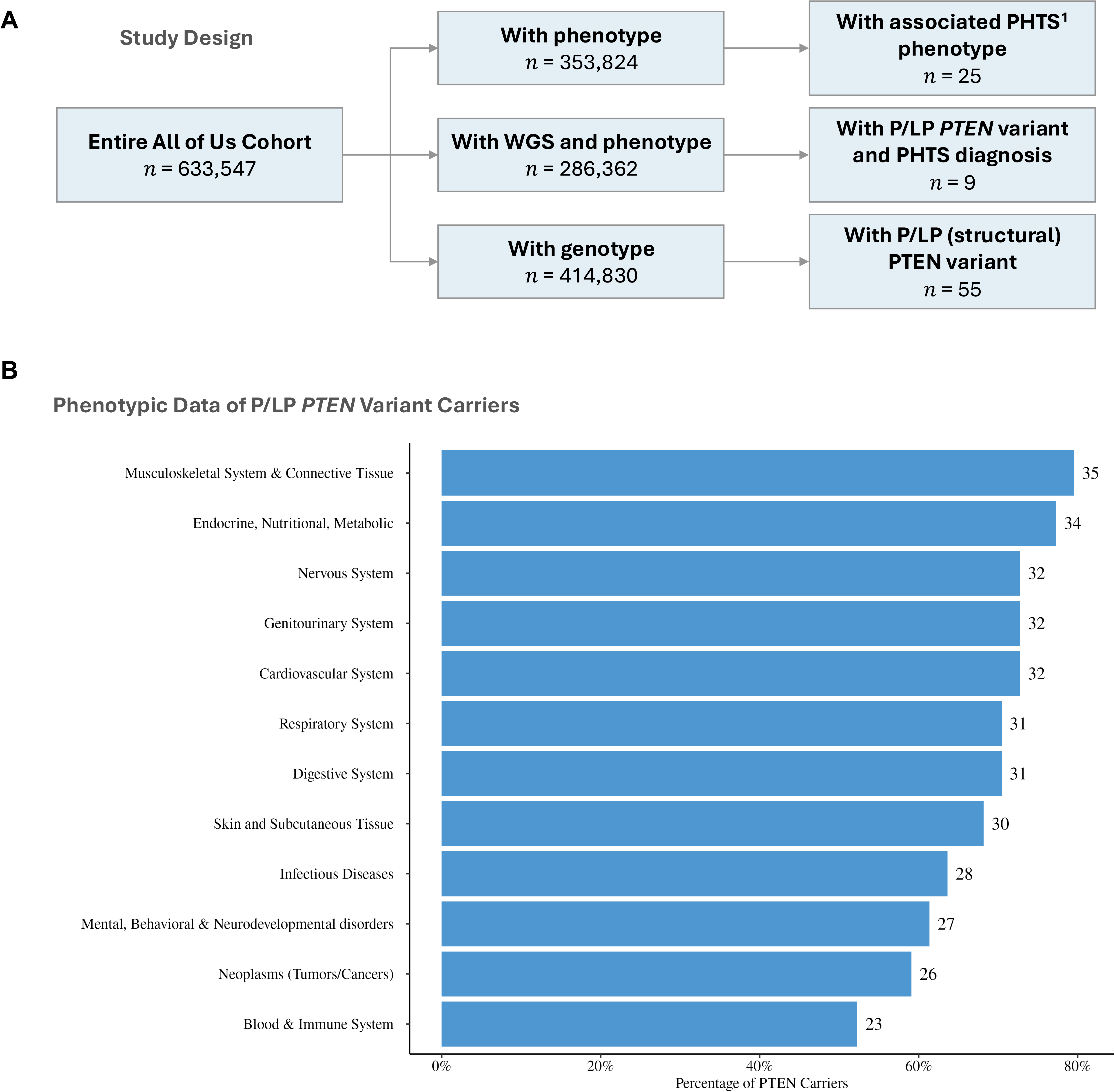
Study Design and Phenotypic Characteristics of Germline *PTEN* Variant Carriers. Panel A, ^1^PHTS phenotype include: *PTEN* hamartoma tumor syndrome, Cowden syndrome, Lhermitte-Duclos disease, Bannayan syndrome, Proteus syndrome and Proteus like syndrome. Panel B, phenotypes have been grouped by 22 ICD-10 code chapters, each representing a broad category of conditions and diseases.

### Comparison to Participants with Germline Variants in Other Cancer-related Genes

In a recent study, we analyzed the AoU database to identify the prevalence of germline P/LP variants in 85 cancer susceptibility genes.^15^ We identified 3,454 unique germline P/LP variants across 77 transcripts and 72 genes, including *PTEN*. Our analysis revealed that 20,968 participants had germline P/LP variants, of which 55 were in individuals with P/LP variants in *PTEN*. Importantly, no participant with germline P/LP *PTEN* variant harbored other germline P/LP cancer susceptibility gene variants. To better characterize the phenotypic spectrum, particularly as related to cancer in a predominantly adult population, we then generated three groups of participants, including those with germline P/LP *PTEN* variants, those with germline P/LP variants in cancer predisposition genes other than *PTEN*, and those without germline P/LP variants in the known cancer predisposition genes (**Table 1**). Importantly, individuals with germline *PTEN* variants had the highest prevalence of cancer compared to the other two groups, including those with germline variants in other cancer predisposition genes (OR, 2.31; 95% CI 1.27-4.14; *P*=0.007). Additionally, the ages at the first cancer diagnosis were younger in participants with germline *PTEN* variants compared to the other two groups (**Table 1**). We plotted the cumulative probability of cancer at age of initial diagnosis comparing the three distinct variant groups to show a median age of onset at 48 years (range 8 - 67 years) for *PTEN* variant carriers, 59 years (range 11 - 99 years) for carriers of other P/LP cancer-related gene variants, and 61 years (range 3 - 103 years) for non-carriers (**Fig. 2**).

**Figure 2:**
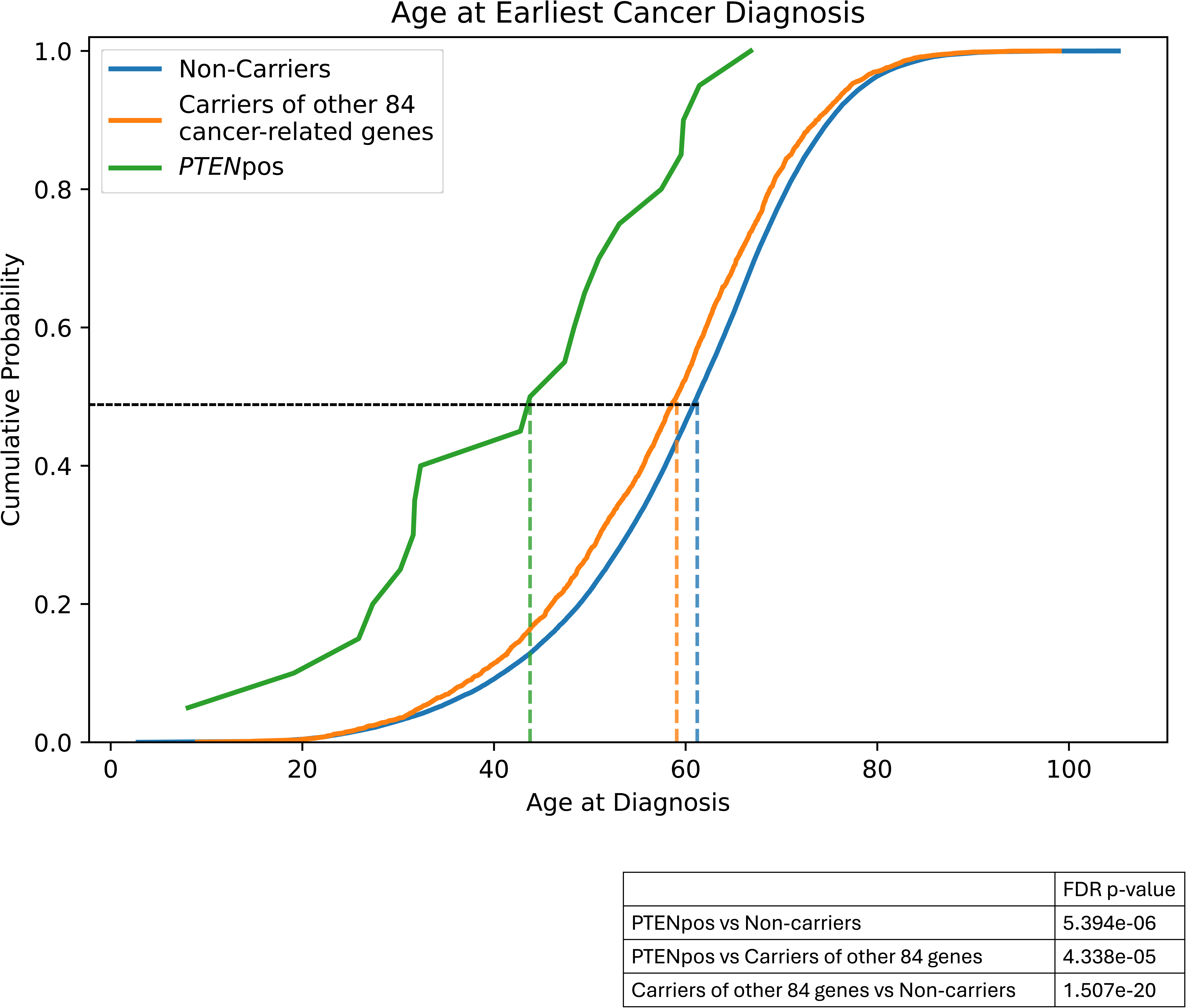
Age at the Earliest Cancer Diagnosis. X-axis represents age at diagnosis in years. Non-carrier, participants without germline P/LP variants in the known cancer predisposition genes, including *PTEN*; Other 84 genes, participants with germline P/LP variants in cancer predisposition genes other than *PTEN*; PTEN, participants with germline P/LP *PTEN* variants, including the two participants with structural variants.

**Table 1:** Comparison of Demographic and Cancer-Related Characteristics by Variant Carrier Status.

| Variable | Carriers of P/LP *PTEN* Variant<sup>1</sup><br>N = 55 | Carriers of Other 84 P/LP Cancer Gene Variants<br>N = 20,915 | Non-Carriers of 85 P/LP Cancer Gene Variants<br>N = 393,860 | p-value<sup>3</sup> |
| --- | --- | --- | --- | --- |
| **Sex<sup>2</sup>** |  |  |  | <0.001 |
| Female | 31 (59.6%) | 12,736 (61.5%) | 237,304 (60.9%) |  |
| Male | 21 (40.4%) | 7,973 (38.5%) | 152,380 (39.1%) |  |
| **Race** |  |  |  | <0.001 |
| American Indian or Alaska Native | 0 (0%) | 239 (1.1%) | 5,515 (1.4%) |  |
| Asian | * | 435 (2.1%) | 13,051 (3.3%) |  |
| Black or African American | * | 2,543 (12%) | 69,989 (17.8%) |  |
| Middle Eastern or North African | 0 (0%) | 95 (0.5%) | 2,256 (0.6%) |  |
| More than one population | * | 896 (4.3%) | 16,920 (4.3%) |  |
| Native Hawaiian or Other Pacific Islander | 0 (0%) | 21 (0.1%) | 435 (0.1%) |  |
| Other/Unspecified | * | 3,911 (19%) | 74,508 (18.9%) |  |
| White | 36 (65%) | 12,775 (61%) | 211,186 (53.6%) |  |
| **Cancer Diagnosis<sup>4</sup>** |  |  |  | <0.001 |
| No | 26 (56.7%) | 11,058 (75.0%) | 220,688 (81.3%) |  |
| Yes | 20 (43.5%) | 3,688 (25.0%) | 50,882 (18.7%) |  |
| **Age at First Cancer Diagnosis<sup>5</sup>** | 46 (8 – 67) | 59 (9 – 99) | 61 (3 – 105) | <0.001 |

<sup>1</sup> Includes 2 individuals with structural variant.
<sup>2</sup> We only report sex count for male and female.
<sup>3</sup> p-values calculated using Kruskal-Wallis test for age and Chi-Squared test for others.
<sup>4</sup> Restricted to participants who have both srWGS and EHR data, therefore the sum may not equal N.
<sup>5</sup> Refers to first of any kind of cancer; n (%), Median (Min – Max).

### Phenotype Enrichment Analysis

Intrigued by this observation, we then sought to perform phenotype enrichment analysis between participants with germline P/LP *PTEN* variants versus those who are wildtype for *PTEN*, including those who have germline variants in the 84 other cancer susceptibility genes. We identified known associations with the PHTS phenotype, including neurocutaneous syndrome, congenital anomalies of peripheral blood vessels, congenital hamartoma, goiter, gastrointestinal polyps, endocrine disorders, benign neoplasms, and others (**Fig. 3A**).

**Figure 3:**
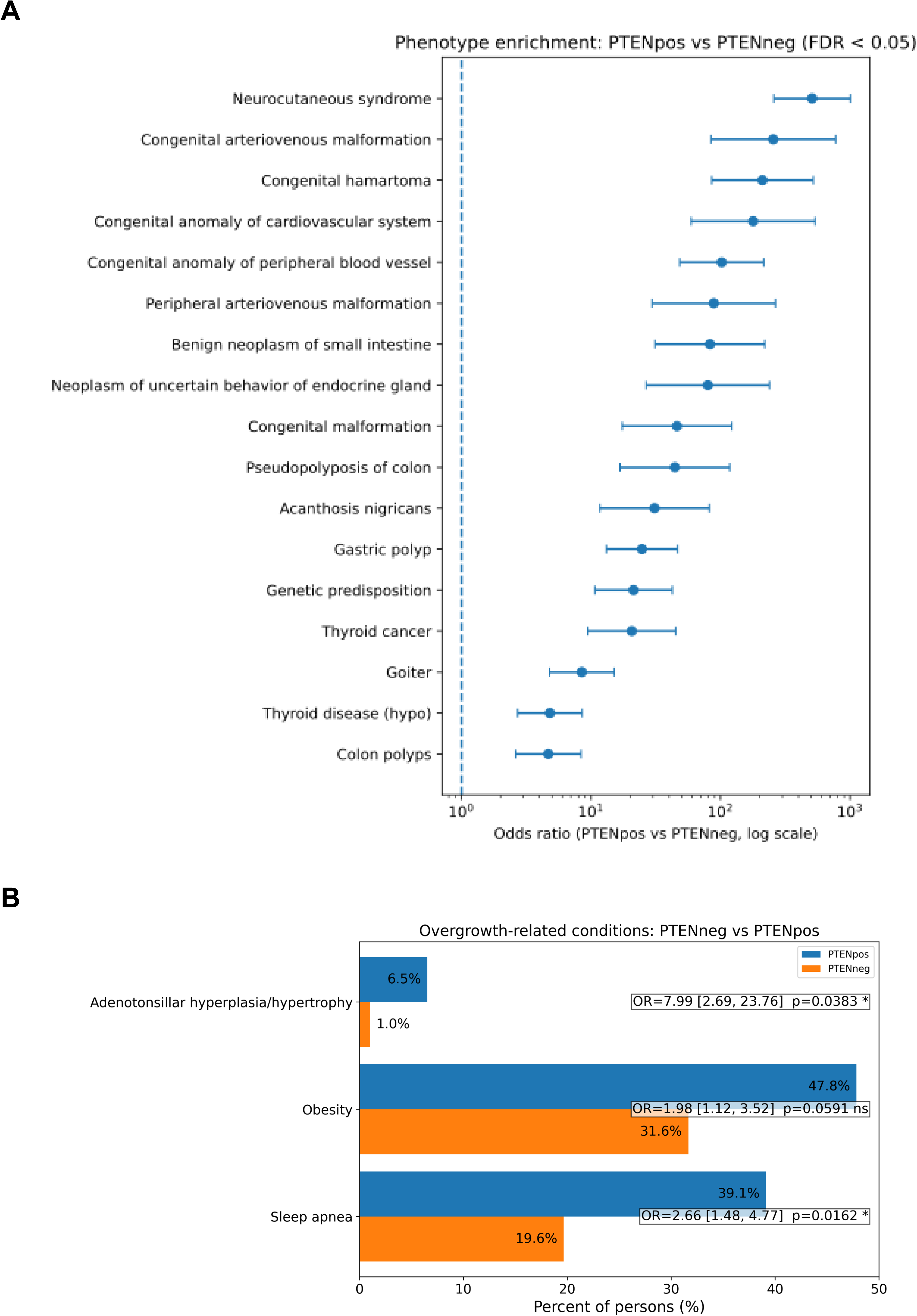
Phenotype Enrichment in Germline *PTEN* Variant Carriers. Panel A, Phenotype enrichment in participants with germline *PTEN* variants versus those who are wildtype for *PTEN*. Panel B, Enrichment focused on overgrowth-related conditions.

Importantly, these phenotypes centered around the overgrowth features of the syndrome. Other previously unrecognized associations include acanthosis nigricans. In an independent targeted analysis, we focused on other overgrowth phenotypes, including hyperplasia of the adenoids and/or tonsils, sleep apnea, and obesity. We identified significant enrichment in adenotonsillar hyperplasia/hypertrophy and sleep apnea in participants with germline *PTEN* variants compared to those without germline *PTEN* variants (**Fig. 3B**). While obesity was not statistically significant, we observed that extreme obesity with alveolar hypoventilation was significantly enriched in participants with germline *PTEN* variants compared to those without (OR, 19.69; 95% CI 6.07-49.92; *P*<0.0001).

### Participants with *PTEN* Variants of Uncertain Significance

Because variants of uncertain significance (VUS) pose a challenge clinically and especially for patient management, we next sought to characterize key features in participants from AoU with *PTEN* VUS. We identified 342 individuals with germline *PTEN* VUS (**Supplementary Table 1**). Notably, compared to participants with P/LP germline *PTEN* variants, carriers of *PTEN* VUS had a lower prevalence of cancer diagnoses (OR, 0.27; 95% CI 0.14-0.53; *P*<0.001), and an older age at first cancer diagnosis (61 years versus 48 years, *P*<0.001) (**Supplementary Table 1** and **Supplementary Fig. 1A**). Similarly to carriers of P/LP germline *PTEN* variants, we observe an overrepresentation of broad phenotypes associated with the musculoskeletal system and connective tissues, in addition to endocrine, digestive, genitourinary, and other phenotypes (**Supplementary Fig. 1B**). However, phenotype enrichment analysis of carriers of germline P/LP *PTEN* variants versus those with VUS revealed an overrepresentation of goiter in the former group (OR, 5.8; 95% CI 2.9-11.4; *P*<0.001).

### Comparison with Cleveland Clinic Patient Series

As related to the *PTEN* genotype, we next sought to investigate the *PTEN* genotype spectrum in the 55 participants with 36 unique single nucleotide variants (SNV) and indels (**Supplementary Table 2**). The most predominant variant types were missense variants (46%), followed by nonsense variants (32%), and frameshift deletions or insertions (22%). We then compared these results to the variant spectrum of 487 individuals with germline P/LP *PTEN* variants from Cleveland Clinic PTEN Multidisciplinary Clinic (**Fig. 4**). In the latter series, the most predominant variant types were missense variants (42%), followed by nonsense variants (27%), and frameshift deletions or insertions (27%). Both series of participants showed an enrichment of hotspot germline *PTEN* variants c.388C>T, p.R130X and c.1003C>T, R335X.

**Figure 4:**
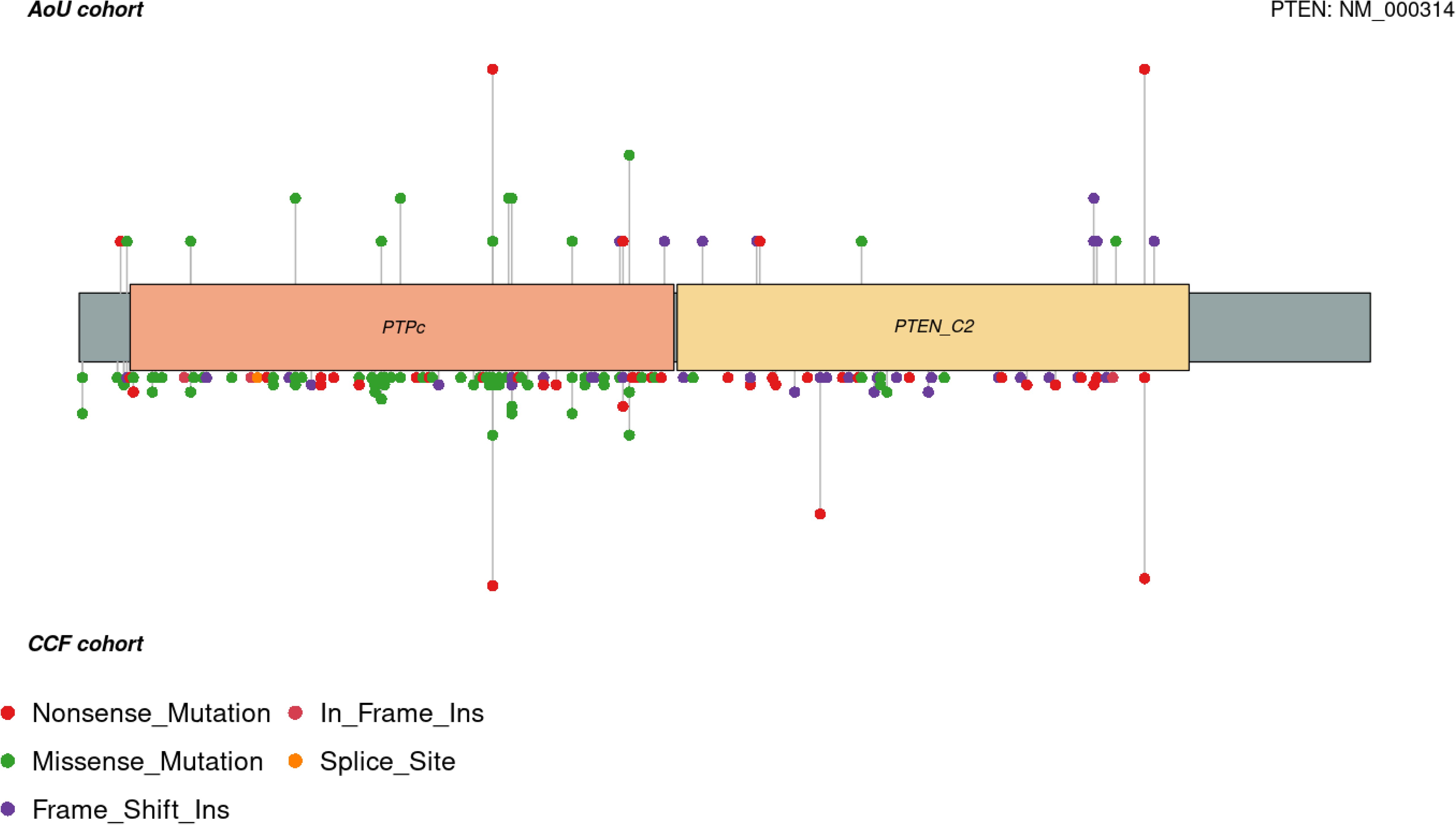
Lollipop Plot of Germline *PTEN* Variants. Lollipop heights were normalized in order not to reveal participant count per variant. Plots do not include structural variants. PTPc, protein tyrosine phosphatase catalytic domain; PTEN_C2, C2 domain of the tumor suppressor protein PTEN.

Finally, we focused on cancer phenotypes, which are well-characterized in both participant series from AoU and the PTEN Multidisciplinary Clinic. This analysis included participants with P/LP germline *PTEN* variants, including SNV, indels, and structural variants. In the AoU dataset, 20 of the 46 (43.5%) participants with available EHR data had at least one cancer diagnosis.

The most predominant cancers were those of the breast (40%), followed by thyroid (35%), endometrial (20%), skin basal cell carcinoma (15%), and other unspecified cancers (15%). Of those with a cancer diagnosis, 13 (65%) of the participants had SMN. In the Cleveland Clinic series, 215 of the 514 (41.8%) participants had at least one cancer diagnosis. Breast (20%), thyroid (14%), endometrial (7%), kidney (6%), and non-melanoma skin (4%) cancers were the most overrepresented cancers. In this series, 98 (45.6%) of participants with cancer had SMN.

## Discussion

In this study, we leveraged the scale and diversity of the AoU research program to characterize the genotype and phenotype spectrum of PHTS in an unselected population-based setting and compared these findings with individuals from a specialized clinical cohort. Our results demonstrate that individuals with PHTS may present differently in a population-based setting, and that many carriers of germline P/LP *PTEN* variants remain undiagnosed, underscoring the underestimated burden of disease in the general population. For example, while macrocephaly (head circumference greater than the 97^th^ percentile for age) is an important and highly prevalent component manifestation of PHTS,^16, 17^ it was not reported in any of the AoU research participants with germline *PTEN* variants. Importantly, until recently, the true prevalence of PHTS was unknown and was based on a study conducted in the Netherlands that estimated a 1:200,000 prevalence of CS in the Dutch population.^14^ Here, we identify a prevalence of 55/414830 or ∼1/7500 individuals, which is about 26 folds higher and consistent with estimates from a study focusing on thyroid cancer in PHTS and other thyroid cancer-associated syndromes.^13^

Our comparison of AoU participants with germline *PTEN* variants to individuals with germline variants in the other cancer predisposition genes revealed an elevated cancer prevalence associated with PTEN dysfunction. Importantly, we also observed that carriers of P/LP *PTEN* variants developed cancer at significantly younger ages than both non-carriers and carriers of other cancer predisposition variants. Given that inclusion criteria are identical for all AoU participants, this reinforces prior observations of elevated lifetime cancer risks in PHTS and highlights the importance for earlier, gene-informed surveillance strategies in this population.^3, 6, 7^ Notably, while AoU participants with *PTEN* variants also exhibited enrichment for phenotypes classically associated with PHTS, including endocrine disorders, vascular anomalies, and gastrointestinal polyps, our enrichment analysis revealed under-recognized associations such as vitamin B deficiency, fibromyalgia, and seizure disorders, warranting further investigation into their potential links with PTEN dysfunction.

Comparison of *PTEN* variant spectra across AoU and the Cleveland Clinic PTEN Multidisciplinary Clinic series revealed similar distributions of missense, nonsense, and frameshift variants, including recurrent hotspot variants such as p.R130X and p.R335X. This convergence suggests that while the molecular spectrum of *PTEN* pathogenic variants is consistent across settings, ascertainment biases shape the clinical profiles of identified carriers. Specifically, AoU participants represent a broader and more heterogeneous population, capturing phenotypes less commonly observed in referral-based cohorts. These findings support the utility of large-scale, population-based sequencing efforts to expand our understanding of the full phenotypic spectrum of PHTS beyond cancer- and NDD-enriched settings.

Our study has important clinical and research implications. First, the identification of undiagnosed carriers within AoU validates that PHTS is indeed underdiagnosed and its prevalence underestimated.^13^ Second, the observation of non-component phenotypes through unbiased enrichment analyses raises the possibility that germline *PTEN* variants contribute to a wider range of systemic manifestations than currently recognized, suggesting opportunities for validation studies to delineate these associations. Finally, our data highlight the need for optimizing the identification of PHTS in community settings, particularly in individuals unaffected by cancer and/or NDD.

This study had limitations. Phenotypic manifestations were not uniformly reported between patients from the *PTEN* Multidisciplinary Clinic and Center of Excellence at the Cleveland Clinic and participants from the AoU research program. This precluded us from performing in-depth comparisons and phenotype enrichment analysis beyond well-characterized cancer phenotypes. Relatedly, the AoU cohort lacked detailed information about classical PHTS-associated phenotypes, such as NDD. Conversely, while the CCF series included pediatric and adult patients, including those with NDD across the lifespan, it lacked granular information regarding non-traditional manifestations, such as obesity.

Overall, our study demonstrates the value of population-scale genomic and phenotypic resources for uncovering the true prevalence and breadth of relatively rare syndromes, such as *PTEN*-related disorders. By studying PHTS within an unselected population, we show that its clinical manifestations extend beyond those captured in specialized clinical cohorts and reaffirm its role as a high-penetrance cancer predisposition syndrome with earlier age of onset compared to other heritable cancer syndromes. Future work integrating longitudinal phenotypic data, and exploration of gene–environment interactions will be essential to refining precision surveillance and management strategies for individuals with PHTS.

## Methods

### Research Participants from All of Us Research Program

We analyzed the controlled tier Curated Data Repository (CDR) version 8 dataset of the All of Us (AoU) research program to investigate the phenotypes and genotypes of individuals with *PTEN*-related disorders. By design, the AoU dataset is a diverse, unselected and large-scale dataset that could be representative of the US population across dimensions such as race, ethnic background, sex, geographic region, socioeconomic background, and health status.^18^ As of the cut-off date of October 1, 2023, the program has enrolled 633,547 participants, out of which 353,834 have phenotypic information and 414,830 have short-read whole genome sequencing (srWGS) data. For the purposes of our analysis, we used the ClinVar callset in the hail MatrixTable format with multiallelic sites split into separate records. The AoU ClinVar callset includes variants reported in the ClinVar database, not limited to pathogenic and/or likely pathogenic variants (P/LP), with a total of 2,180,727 single nucleotide variants (SNVs) and indels, with multiallelic sites split into separate records.

The AOU database contains multiple transcripts for each gene. For our analysis, we only used transcripts that were either classified as Matched Annotation from NCBI and EMBL-EBI (MANE) or MANE Plus Clinical.^19^ The following criteria was used to identify P/LP variants. First, we removed any variant whose consequences is annotated as ‘downstream_gene_variant’ or ‘upstream_gene_variant’. Next, any variant labelled as P/LP but also included other ambiguous annotations such as risk factor, uncertain significance or likely allele were manually verified in the ClinVar website.

We identified participants with PHTS associated conditions as those with the following reported conditions in their EHR data: *PTEN* hamartoma tumor syndrome, Cowden syndrome, Lhermitte-Duclos disease, Bannayan syndrome, Proteus syndrome and Proteus like syndrome.

### Phenotype Data Extraction from AoU

The AoU phenotypic data includes different data types collected from participants including data directly collected by the program (e.g., demographics, surveys and physical measurements), data shared through external electronic health records (EHRs), data from wearable devices, and data from biospecimens. The AoU research program uses the Observational Medical Outcomes Partnership (OMOP) Common Data Model (CDM) to store and standardize participants information provided via surveys, physical measurements and EHRs. All data collected are expressed as “concepts” in OMOP. The EHR data includes reported conditions in several EHR codes including SNOMED, ICD10CM, ICD9CM, Nebraska Lexicon, CIEL, and others. To identify relevant phenotypes, we mapped the documented conditions to ICD10CM. Each condition was assigned to one of 22 ICD-10 chapters, including “*Infectious Diseases,” “Neoplasms (Tumors/Cancers),” “Blood & Immune System,” “Endocrine, Nutritional, Metabolic,” “Mental & Behavioral Health,” “Nervous System,” “Eye and Adnexa,” “Ear and Mastoid Process,” “Cardiovascular System,” “Respiratory System,” “Digestive System,” “Skin and Subcutaneous Tissue,” “Musculoskeletal System & Connective Tissue,” “Genitourinary System,”* and *“Congenital Malformations.”* We excluded conditions categorized under the chapters *“Pregnancy, Childbirth, Puerperium,” “Symptoms & Abnormal Clinical Findings,” “Injury & Poisoning,” “External Causes of Morbidity,”* and *“Health Services / Social Circumstances.”* These grouped conditions formed the basis for subsequent phenotype prevalence analyses except when otherwise stated.

### Phenotype Enrichment Analysis

We utilized the standard concept names (grouped manually into related conditions) associated with each phenotype as recorded in the AoU database. We used Fisher’s exact test to calculate odds ratios, 95% confidence intervals, and p-values. To account for multiple hypothesis testing, p-values were adjusted using the false discovery rate (FDR) method. Phenotypes enriched in the *PTEN* cohort were filtered to those with significant p-values (*P*<0.05). Filtration of prioritized phenotypes shown in Figure 2 included the following criteria: (1) Requiring at least two *PTEN* variant positive (*PTEN*pos) individuals with the phenotype so ORs are not driven by singletons; (2) Requiring at least 50 individuals (*PTEN*pos plus *PTEN* variant negative [*PTEN*neg]) with the phenotype, so the result is not dominated by rare phenotypes; (3) Requiring a minimum of 10 *PTEN*neg individuals with the phenotype to avoid inflated or infinite ORs and finally (4) Keeping only phenotypes with a maximum 95% CI span (CI_hi/CI_lo) of 25.

### Research Participants from the Cleveland Clinic

Research participants from the Cleveland Clinic were prospectively accrued from September 1, 2005, through January 6, 2022, as part of a prospective follow-up study approved by the Cleveland Clinic institutional review board (IRB protocol 8458).^7^ The study was conducted in agreement with the principles of the Declaration of Helsinki. All participants provided informed written consent to participate. Participants were evaluated at the *PTEN* Multidisciplinary Clinic and Center of Excellence at the Cleveland Clinic (Cleveland, Ohio, USA). This study included both pediatric and adult patients with PHTS accrued from community and academic medical centers throughout North and South America, Europe, Australia, and Asia. Reported cancer diagnoses were documented through pathology reports or verified cancer genetics visits.

Baseline information including any cancer history was recorded at the time of consent. Between July 2021 and July 2022, we obtained updated phenotypic information in those who had not routinely seen us in genetics clinic within 3 years.^7, 20^ We reviewed cancer-related health records of patients with PHTS internal to the Cleveland Clinic health system. For the purposes of this study, we prioritized individuals with confirmed P/LP germline *PTEN* variants. *PTEN* variant classifications were ascertained by clinical genetic testing reports where available, ClinVar database classifications, and/or the ClinGen gene-specific criteria for *PTEN* variant curation.^21^

### Statistical Analysis

All statistical analyses were conducted using R studio (version 2024.04.0 Build 735) and Python (3.10.16) programming within the AoU Research Workbench. We analyzed the demographic breakdown of our study cohorts, looking at sex, race, cancer diagnosis status (yes/no), and age at first cancer diagnosis. We utilized the Kruskal-Wallace test to compare age at first cancer diagnosis across cohorts and Mann-Whitney U test for pairwise analyses of age at first diagnosis. We implemented the Chi-Squared test to compare sex, race, and cancer diagnosis breakdowns. A significance threshold of *P*<0.05 was used throughout the study.

## Data Availability

All data produced in the present work are contained in the manuscript.

## Acknowledgments

We are sincerely indebted to the generosity of the families and patients in PTEN clinics across the United States who contributed their time and effort to this study. We thank the clinical research team at the *PTEN* Multidisciplinary Clinic and Center of Excellence at the Cleveland Clinic for administrative support. We gratefully acknowledge All of Us participants for their contributions, without whom this research would not have been possible. We also thank the National Institutes of Health’s All of Us Research Program for making available the participant data examined in this study. This work has been partly funded by the National Institute of Child Health and Human Development (R01HD105049 to Y.N.).

## Data and code availability

All of Us (AoU) data analyses were performed using the AoU Controlled Tier Dataset v8 dataset and executed using the AoU Research Workbench. All other raw data are available from the corresponding author on reasonable request. All other bioinformatics and statistical tools are publicly available and are described above.

## Author contributions

Y.N. conceptualized and designed the project. C.B., G.I., and L.Y. collected the data. C.B., G.I., L.L., and L.Y. performed the data analyses. L.Y., G.I., and Y.N. interpreted the data. G.I. and C.B. performed primary bioinformatic and statistical analysis of the data. G.I., C.B., and L.Y. drafted the original version of the manuscript. Y.N. and L.Y. supervised the project. All authors read, critically revised, and approved the final version of the manuscript.

